# Medical laboratory waste generation rate, management practices and associated factors in Addis Ababa, Ethiopia

**DOI:** 10.1101/2021.08.17.21262112

**Authors:** Salem Endris, Zemenu Tamir, Abay Sisay

## Abstract

**Background:** Biomedical wastes (BMW) generated from medical laboratories are hazardous and can be deleterious to humans and the environment. Highly infectious types of biomedical wastes are commonly generated at an unacceptably high rate from health laboratories of developing countries with a poor management system like Ethiopia. This study was aimed to evaluate the generation rate of biomedical wastes, management practices and associated factors among public healthcare medical laboratories in Addis Ababa, Ethiopia.

**Materials and methods:** Health institution based cross-sectional study was conducted from July 13 to September 25, 2020 in 6 hospital laboratories and 20 health center laboratories in Addis Ababa, Ethiopia.Data on socio-demographic characteristics, knowledge and practice of biomedical wast managment of the health facilities, Biomedical waste generation rate were collected using data collection tools. Data were managed using SPSS version 20 software. Descriptive statistcs,Pearson correlation, linear and logistic regression analysis were computed to identify indepedent predictors of the dependent variable. Odds ratio with 95% confidence interval was used to determine the strength of association.

**Results:** The finding revealed that the mean ± SD of the daily generation rate of biomedical wastes was 4.9 ± 3.13 kg/day per medical laboratory. Nineteen (74.3%) medical laboratories had a proper practice of biomedical waste management which is significantly associated with knowledge of professsionals on biomedical waste management policies and guidelines, availability of separate financial sources for biomedical waste management and training level of professionals.

**Conclusion:** The study showed high generation of biomedical waste from medical laboratories in public healthcare of Addis Ababa,Ethiopia. Near two-thirds of health facilities had a proper practice of waste segregation, collection, storage, and treatment of biomedical wastes generated from their laboratory. However, there was a poor practice of transportation and disposal. Hence, due attention and practicing as per the current national guidelines of biomedical waste management is recoomended.

## Introduction

Biomedical waste represents all wastes generated by health care establishments, research facilities, and clinical laboratories during the diagnosis, treatment, or immunization of human beings or in related research activities that present a potential risk in human health than any other waste[1,2]. Public health and diagnostic laboratories generate a sizable amount of biomedical waste (BMW). With the emerging need for medical service and technology, Medical Laboratories generate significant volumes of hazardious wastes that need special packaging and or handling as well as treatment procedures[3,4]. Higher load of chemical wastes, clinical glass, culture plates, stock cultures, highly infectious wastes in large quantities, and some radioactive wastes are generated from laboratories[1,5].

Inappropriate management of these hazardous biomedical wastes results in exposing staffs in health care facilities, patients, waste collectors and the public in general to infections pathogens[3]. Infectious healthcare wastes can transmit more than 30 dangerous bloodborne pathogens, but the pathogens that are found to be significant are hepatitis B, hepatitis C, and Human immune deficiency virus (HIV). It is estimated that in the year 2000, needle stick injuries caused 21 million hepatitis B, 2million hepatitis C, and 260,000 HIV infections[6,7]. Proper management of infectious biomedical wastes shuld not be optional rather mandatory[8]. Hence, appropriate healthcare waste management with its vital steps; control of generation, segregation, collection, storage, transport, treatment, and disposal in a manner that follows best principles of public health, economics, engineering, conservation, aesthetics, and other environmental considerations is very crucial [2,6].

Hazardous wastes generated in healthcare facilities were showen as unacceptably high which ranges from 21 to 70% and the waste management practice in the country is indicated as poor[6]. It was shown that absence of policy or regulations regarding biomedical waste management, lack of awareness and inadequate training of professionals, limited human resource and financial scarcity in many health facilities are related to poor management of wastes and lack of provision of protective measures[6,9-11]. However, the generation rate and management practice of wastes among medical laboratories in Ethiopia which are important sources of infectious wastes is not well documented yet.

Hence, this study was conducted to asess the generation rate and management system of biomedical wastes of medical laboratories and its associated factors in Addis Ababa, Ethiopia which helps to identify factors affecting BMWM, initiate stakeholders to amend or develop more extensive policies and strategies and alarm laboratory managers to give attention to BMWM.

## Materials and Methods

### Study design, Setting, and participants

Health institution based cross-sectional study was conducted from July 13 to September 25, 2020 in selected public healthcare facility laboratories (Health centers and Hospitals) in Addis Ababa, Ethiopia. Addis Ababa is the largest and capital city of Ethiopia with an estimated population of 6.5 million, holding 527 square kilometers divided into 10 sub-cities[12].There are 99 health centers and 6 public health hospitals in Addis Ababa under Addis Ababa city administration. Each Hospital and health center serves to 1 – 1.5 million and 40,000 people, respectively[13,14].

A total of 26 Public health facilities were included in the study and categorized based on the sub-city where they are found. All (6) public hospitals and twenty health centers which are selected by a simple random sampling method were included. The number of health centers to be included from each sub-city was determined by proportionate allocation to their numbers. Convenience sampling method was used to select study participants (medical laboratory professionals and waste handlers) from each public health facility. During the data collection period, 25 -26 laboratory professionals and 4 waste handlers from each public hospital; and 7 laboratory professionals and 2 waste handlers from each health center who were available and volunteer to participate in the study were enrolled. Medical laboratory managers and quality officers were selected purposively and included in the study.

### Data collection Procedure

Primary data was collected using a data collection tool prepared for this study purpose and recorded. Daily measurement of solid biomedical wastes generated from each laboratory was conducted using a daily precaliberated weighing scale(kg/day) for consecutive seven days. Empty plastic bags with standard color were provided and each was labeled to indicate sample number, date of collection, place of generation, and type of the waste.

Self-administered questionnaire which was developed by reviewing literatures, articles, international and national biomedical waste management guidelines and assessment tools[1-3,10,12,15-18], was used to assess biomedical waste management practices and related factors. The questionnaire consisted of three sections which are socio-demographic characteristics, knowledge of laboratory professionals regarding BMWM, the practice of BMWM of the health facilities, and associated factors. An observational checklist and interview were also used to assess the actual practice of the BMWM system. The data collection tool was pretested in 5% of the sample before commencement of the actual data collection. Two trained data collectors collected the data under the supervision of the principal investigator for three months and both Amharic and English languages were used as a medium for data collection.

#### Operational definition of terms

Biomedical waste: a waste generated from medical laboratories during an investigation of body fluids like blood, urine, stool, sputum and other body fluidsswhich could be hazardous or non – hazardous.

Biomedical waste management system: is a system that controls the generation, segregation, collection, storage, transport, treatment, and disposal of biomedical wastes generated from medical laboratories in health care facilities.

Proper practice: is a public health facility that performs more than three components of biomedical waste management system accordingly to national policy and guideline of health care waste management in Ethiopia

Poor practice: means a public health facility that practice less than three components of the biomedical waste management system.

#### Data analysis and interpretation

Data were entered into a statistical package for social science (SPSS) version 20 for analysis. Descriptive statistcs were used to summarize data and presented using tables. Pearson correlation, linear, bivariable, and multivariable logistic regression analysis were computed to identify indepedent predictors of the dependent variable. Odds ratio with 95% confidence interval was used to determine the strength of association.

### Ethiccal consideration

Ethical clearance was obtained from the research ethics review committee of the department of Medical Laboratory Science College of Health Sciences, Addis Ababa University and a letter of request was sent to Addis Ababa Health Bureau. Official letters from Addis Ababa public health research and emergency management directorate were written to six public hospitals and ten sub-cities. Each sub-cities respective health offices wrote permission letters to conduct the study in selected health centers. Informed consent was obtained from each of the study participants after briefing on the purpose and significance of the study. Personal identifiers were removed and only codes were used throughout the study to keep anonymosity of the study participants.

## Results

### Sociodemographic characteristics

In this study, a total of 362 participants (298 medical laboratory professionals and 64 waste handlers) were included from 26 healthcare facilities. Among these, 194 (53.6%) were females. The mean ± SD age of respondents was 30.4 ± 6.63 years.. Regarding the level of education, 212 (58.6%) and 84(23.2%) of study participants were holders of Bachelor’s degree and Diploma respectively. Among all, 152(42%) and 130(35.9%) had work experience of 5 – 10 years and 1 – 5 years respectively [table 1].

**Table 1:**
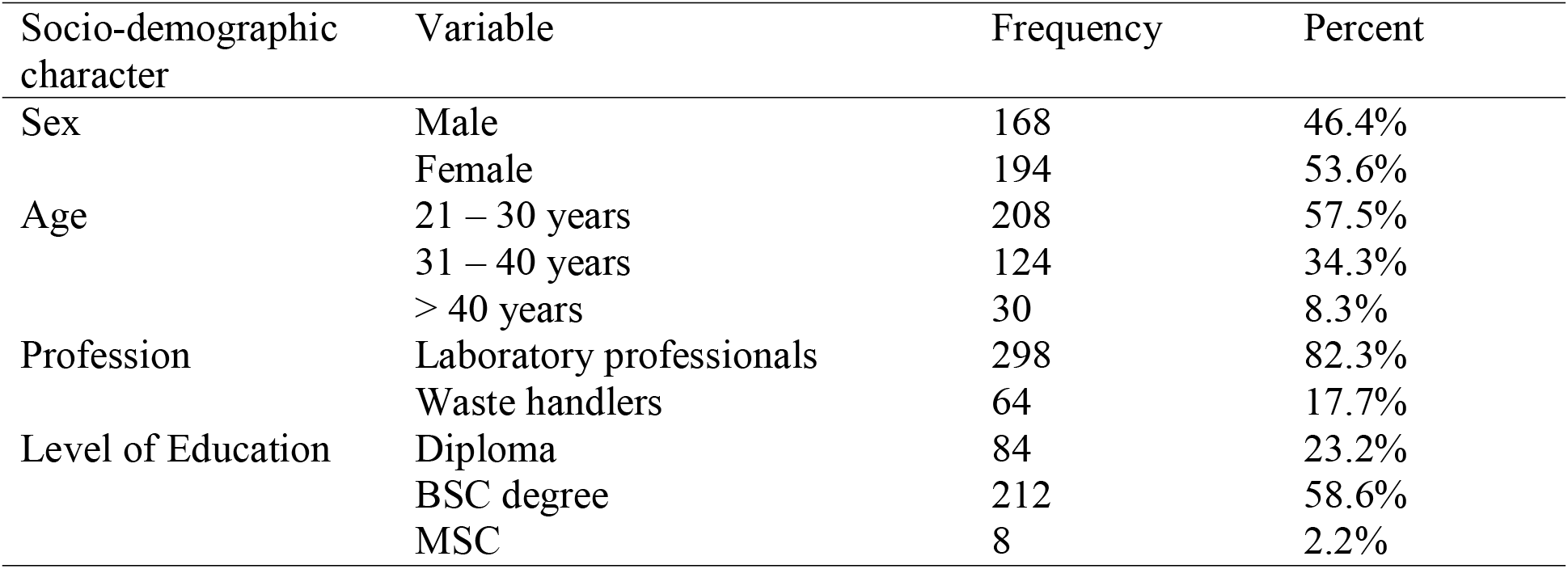
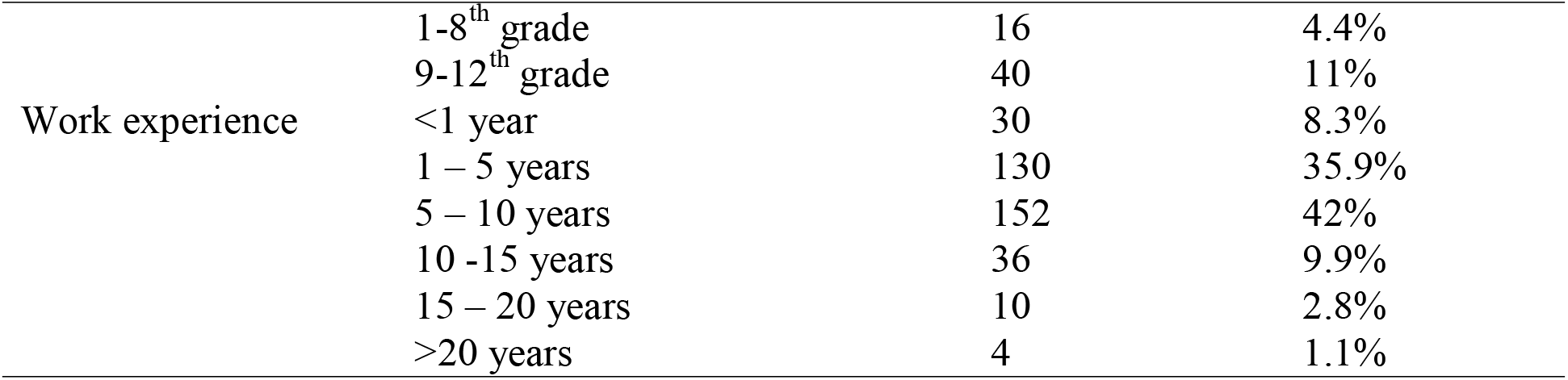
Sociodemographic characteristics of study participants for a study of generation rate and biomedical waste management of medical laboratories and its associated factor at selected public health facilities in Addis Ababa

### Knowledge of study participants about the biomedical waste management

The majority of participated laboratory professionals (63%) and of waste handlers (81.3%) included in this study knew about biomedical waste management policy and guidelines in Ethiopia. Regarding the storage time, 198(54.7%), 74(20.4%), 20(5.5%), and 6(1.7%) knew that the maximum time of storage of biomedical waste in the facility is 12 – 24hrs, less than 12hrs, 24 – 48hrs and greater than 48hrs respectively. Only 40(11%) of study participants explained that incineration is the common mode of treatment of biomedical waste before final disposal, the other 128(35.4%) said that bleaching is a common-mode treatment. More than three fourth (78.5%) of the study participants knew that the safety box has to be disposed of when it is ¾ filled. Half (50.3%) of laboratory professionals and fifty-eight (90.6%) owaste handlers had received training about biomedical waste management, [table 2].

**Table 2:**
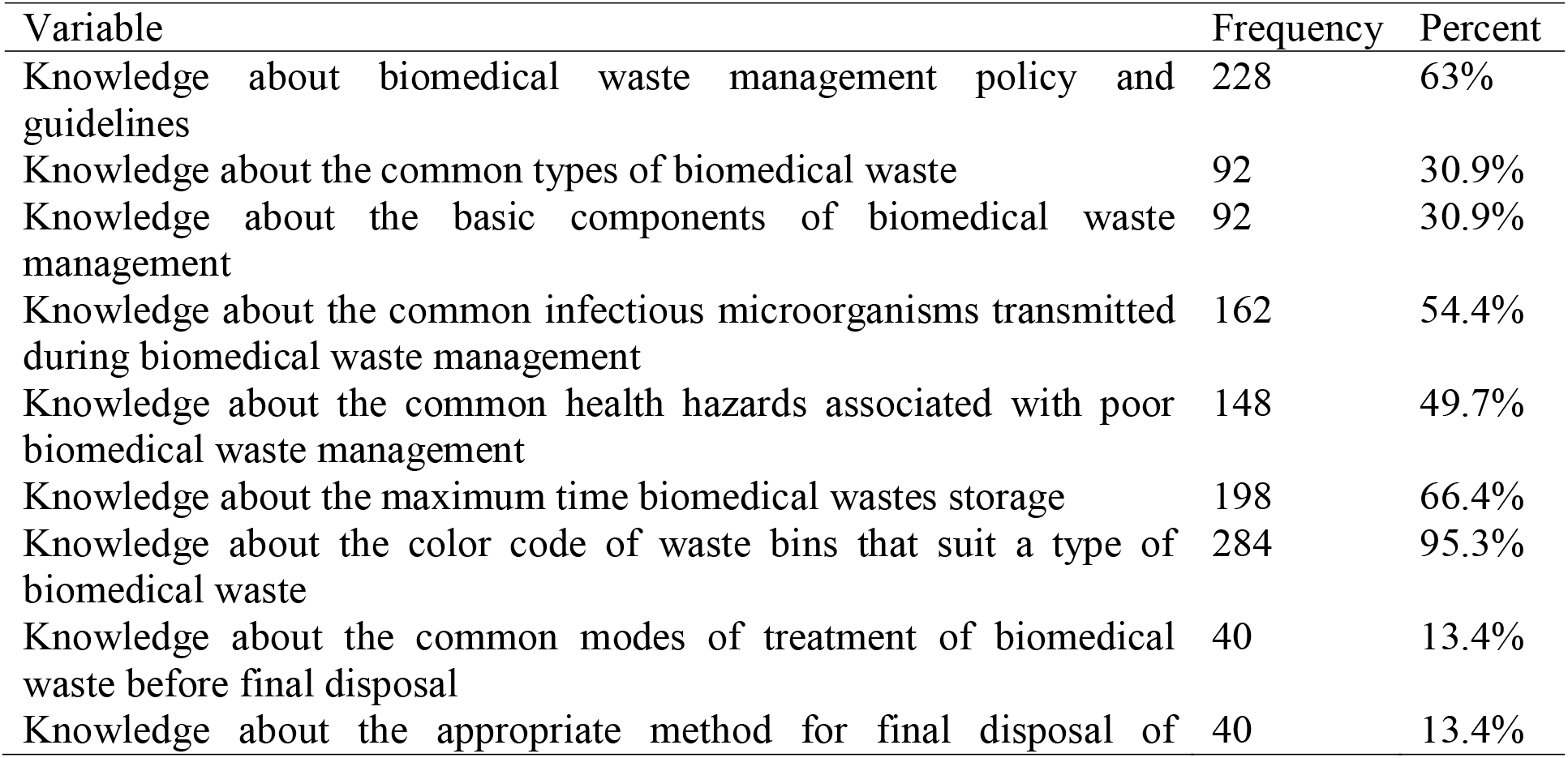
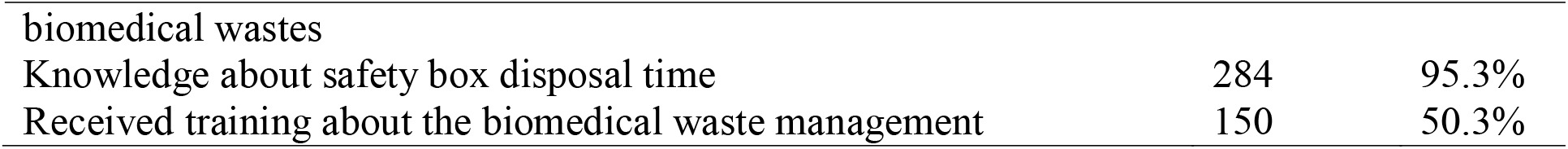
Frequency of laboratory professionals of favorable answers for knowledge item questions for a study of biomedical waste management (n=298)

### Biomedical waste management practice

Two hundred ninety-six (99.3%) medical laboratory professionals replied that there are separate containers for the collection of hazardous and non – hazardous biomedical wastes in their laboratory. Among these, 286(96%) confirmed that they used color coding based segregation whereas the rest 10(3.4%) do not use color coding as segregation and those 264(88.6%) waste containers have a biohazard symbol. Four healthcare facilities (16%) do not follow proper segregation practice. About 56.3%, 28.1%, and 15.6% of waste handlers confirmed that as they are collecting biomedical wastes from the laboratory every 12 hours, 8 hours, and 24 hours respectively. Regarding sharp waste management, 276(92.6%) medical laboratory professionals indicated that safety boxes are available in arm reach places for collection of sharp waste and collected when it is ¾ filled according to 94.6% of participants. Two hundred fifty-six (85.9%) medical laboratory professionals and 63 (96.9%) waste handlers explained that waste holding bags and containers are durable enough. The use of personal protective equipment during waste collection was practiced in all health care facilities. However, (79.2%of medical laboratory professionals and 90.6% waste handlers have confirmed that their facility does provide personal protective equipment. In the contrary, 18.2% of participants have reported as they encountered physical injuries like needle stick injury and sharp injury.

Regarding the precaution and method of transportation of biomedical wastes, 48.3%, 23.5%, 18.1%, and 8.1% of medical laboratory professionals indicated that holding the waste-collecting bags with bare hands, use of wheelbarrows, use of trolley and by the waste container itself are the methods used for transportation of biomedical wastes, respectively. Twenty-six (40.6%) waste handlers also confirmed that there is no separate transportation tool for biomedical wastes in their facility. According to 63% of the study participants, infectious waste collection bins had become covered since the coronavirus pandemic.

Majority of healthcare facilities have storage area far from the storerooms of medical equipment and café as confirmed by 93.8% of waste handlers. According to 78.1% of waste handlers, collected biomedical wastes get stored for less than 12 hours.

About 70.2% of the medical laboratory professionals confirmed that infectious wastes get treated before disposal by using incineration, autoclaving, and chemical disinfection methods. Liquid wastes also get decontaminated first before dumping into running water as 69.6% of medical laboratory professionals explained whereas 11.6% said that it just gets dumped into running water without decontamination. Regarding the waste disposal practice, 38.1%, 18.8% and 12.2% of medical laboratory professionals indicated that open burning pits, landfills, and direct to the municipal waste system are the common types of disposal methods in their facility; whreas 59.4%, 9.4%, and 12.5% of waste handlers replied that treated biomedical wastes get disposed of in open pit, landfills, and municipal waste systems, respectively.

In this study, 67.4% of study participants confirmed that the manager of the medical laboratory in their facility is concerned about biomedical waste management as their routine work. The study also indicated only 18.2% of participants did confirm that there is a separate financial source for biomedical waste management in their institution and 49.7% believed that their institutin legitimately follows the current guidelines of biomedical waste management in Ethiopia. The study also revealed that 69.6% and 6.1% of medical laboratory professionals got vaccinated for both hepatitis B virus and tetanus and only for hepatitis B virus, respectively; whereas 6.6% of them didn’t get the vaccination for both.

### *Waste Generation* in health care facilites

In this study, the mean ±SD of daily solid biomedical waste generation per laboratory in the healthcare facilities was 4.9 ± 3.13 kg/day. Among these, 3.86 ± 2.66 kg/day was a hazardous waste and 1.10 ± 0.853 kg/day was a sharp waste. In average about 112 (± 67)patients get medical laboratory services per day in the sampled facilities. During our onsite observation, we found that slightly higher than half (56%) of the healthcare facilities had a waste storage area whereas eleven (44%) of them had no storage area; most of the facilities used incineration as a method of treatment of biomedical wastes, more specifically, 23(92%) of incinerators were low temperature (made of brick and clay); in 23 (92%) healthcare facilities, waste disposal sites were found far away from any water source; in 21(84%) healthcare facilities, separate landfills are used for waste disposal; more than three fourth of the facilities disinfect liquid waste before disposal, and got collected in a septic tank in 24(96%) facilities. All together, as per the observation, near two-thirds(74.3%) of selected public health facilities properly practice one or more than of the components of the biomedical waste management system, table 3.

**Table 3.** Daily laboratory solid waste generation rate in public health care facilities in Addis Ababa

| Name of health facility | Biomedical waste, kg/day |  |  |  |
| --- | --- | --- | --- | --- |
|  | Total solid waste kg/day | Mean Infectious waste<sup>18</sup> | Mean of sharp waste (MS) | The overall mean of each facility from MI and MS |
| Goro HC | 11.5 | 3.5 | 0.3 | 1.91 |
| Ferency Woreda 01 HC | 3 | 0.76 | 0.23 | 0.49 |
| Entoto kuter 1 HC | 5.5 | 1.25 | 1 | 1.125 |
| Tirunesh Beijing hospital | 9.55 | 2.265 | 0.915 | 1.59 |
| Ras desta hospital | 31 | 10 | 0.33 | 5.17 |
| Mikiland HC | 12.5 | 3.13 | 0.83 | 1.98 |
| Nifas selk woreda 2 HC | 8.5 | 2.5 | 0.33 | 1.41 |
| Arada HC | 7.5 | 1.83 | 0.66 | 1.25 |
| Ghandi memorial hospital | 6 | 1.66 | 0.33 | 0.995 |
| Menelik II hospital | 13.65 | 3.91 | 0.63 | 2.27 |
| Bisrate Gebirale HC | 14.9 | 4.5 | 0.46 | 2.48 |
| Hiwot Amba HC | 6.2 | 1.7 | 0.33 | 1.015 |
| Zewditu memorial hospital | 36 | 10 | 2 | 6 |
| Janmeda HC | 17 | 4.76 | 0.92 | 2.84 |
| Teklehaymanot HC | 16.7 | 4.43 | 1.13 | 2.78 |
| Kolfe HC | 20.1 | 5.33 | 1.36 | 3.34 |
| Yekatit 12 hospital | 10.58 | 3.303 | 1.953 | 2.62 |
| Saris HC | 13.2 | 3.6 | 0.8 | 4.4 |
| Mychew HC | 28.4 | 6.13 | 3.33 | 4.73 |
| Kality HC | 9.5 | 1.66 | 1.5 | 1.58 |
| Addis ketema Woreda 4 HC | 19.6 | 5.3 | 1.8 | 3.55 |
| Addis ketema woreda 7 HC | 7 | 1.25 | 1.08 | 1.16 |
| Entoto No.2 HC | 29 | 6.9 | 2.76 | 4.83 |
| Addis Gebeya HC | 17.2 | 3.6 | 2.13 | 2.86 |
| Semit HC | 9.7 | 2.66 | 0.56 | 1.61 |
| Kasanchis HC | 7.8 | 2.06 | 0.53 | 1.29 |
| Overall mean | 14.8052 | 3.8653 | 1.1004 | 4.947 |
| SD | 8.75676 | 2.6676 | 0.85389 | 3.1348 |

### Factors associated with biomedical waste management

In multivariate regression analysis; age, profession, and level of education showed marginal association with knowledge. Sex, working experience and training found to have a statistically significant association with knowledge; male gender (AOR: 2.771 95% CI (1.164, 6.596)), work experience of 1 – 5 years (AOR: 344 95% CI (19, 6009)) and 5 – 10 years (AOR: 113, 95% CI (7.5, 1683.9)) have an association with knowledge of policies and guidelines regarding biomedical wastes management, common infectious microorganism related to poor waste handling, and common modes of treatment. Having previous training is found to be associated with knowledge about most common infectious microorganisms and the maximum time of waste storage shown as (AOR: 7.29 95% CI (1.57, 33.7 p-value: 0.011)), (AOR: 5.635 95% CI (1.461, 21.730 p-value: 0.012)) respectively. (Table 4).

**Table 4:** Multivariate logistic regression analysis of factors related to knowledge regarding biomedical waste management

| Variable |  | Knowledge status |  | AOR | 95% CI |  | P-value |
| --- | --- | --- | --- | --- | --- | --- | --- |
|  |  | Yes | No |  | Lower | Upper |  |
| Gender | Male | 98 | 70 | 2.771 | 1.164 | 6.596 | .021 |
|  | Female | 64 | 66 | 0.862 | 1 |  |  |
| Work experience | <1 year | 14 | 6 | 2.824 | 2.67 | 4.21 | .974 |
|  | 1-5 years | 68 | 30 | 344.758 | 19.778 | 512.6 | .000 |
|  | 5-10 years | 112 | 22 | 113.050 | 7.590 | 345.6 | .001 |
|  | 10-15 Years | 24 | 10 | 605 | 582 | 690 | .991 |
|  | 15-20 years | 8 | 2 | 174.8 | 164 | 238 | .983 |
|  | >20 years | 2 | 0 | 729.1 | 1 |  |  |
| Training | Yes | 180 | 2 | 7.290 | 1.574 | 33.767 | .011 |
|  | No | 102 | 12 | 0.094 | 1 |  |  |

Similarly, availability of standard operational procedure, providing durable waste holding bags, method of waste transport, treating infectious wastes before disposal, presence of separate financial source, legitimately following the current guideline of biomedical waste management in medical laboratories is strongly associated with knowledge of biomedical waste management system, (Table 5).

**Table 5:**
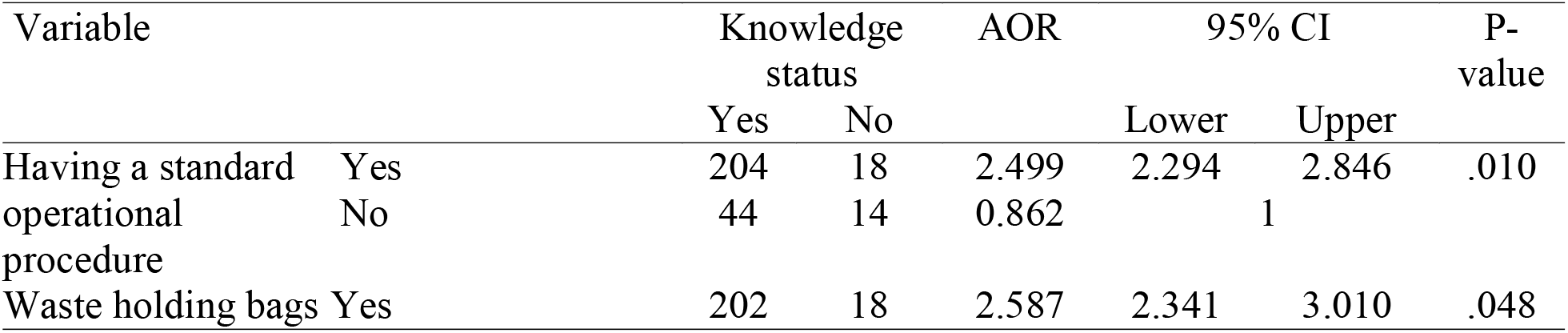
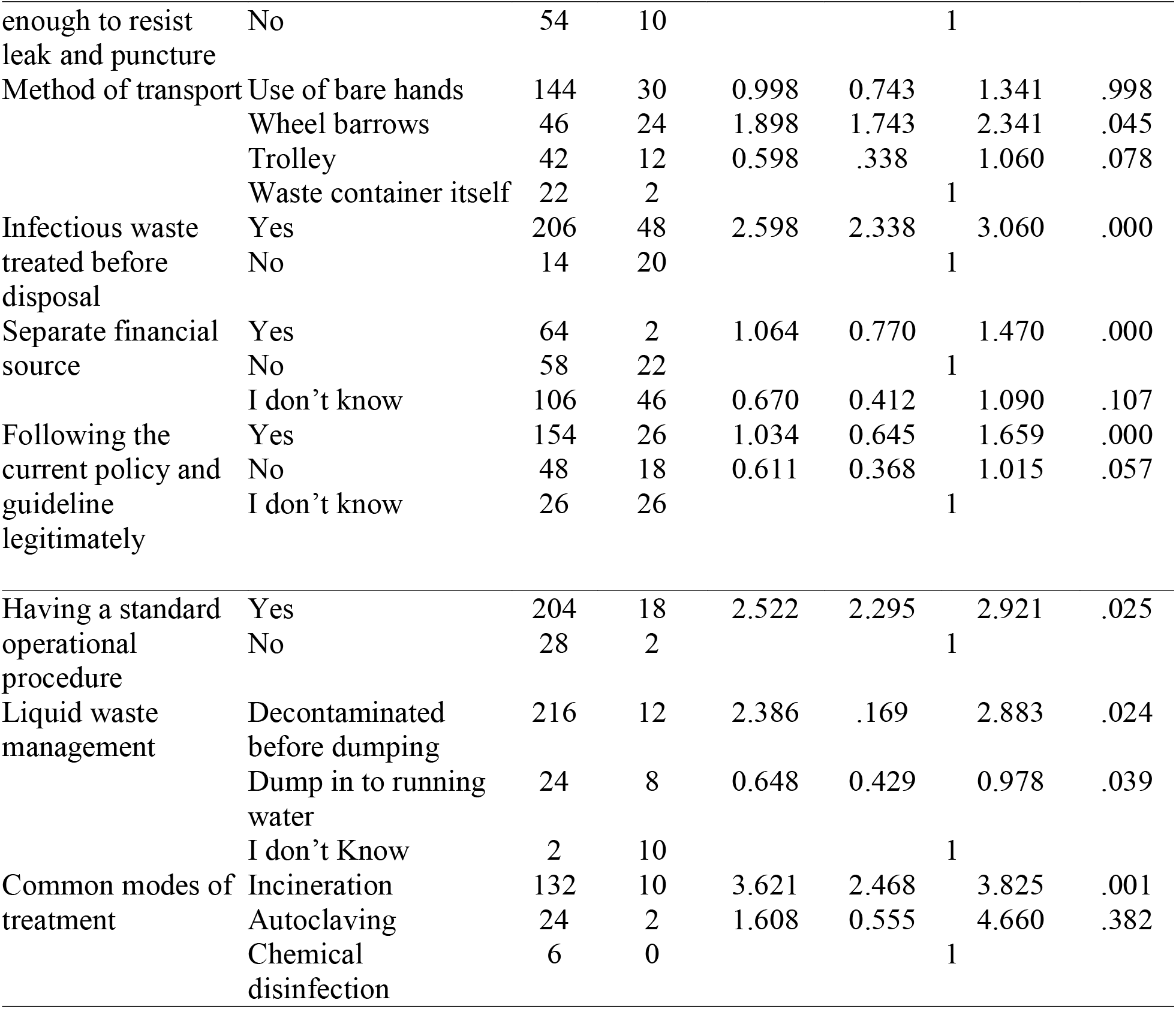
Multivariate logistic regression analysis which shows the association of knowledge with the practice of biomedical waste management.

In linear regression analysis, the number of patient getting medical laboratory service per day showed a statistically significant association with daily total solid waste generation rate per medical laboratory (t=3.032; 95% CI (2.421, 12.999)). table 6.

**Table 6:**
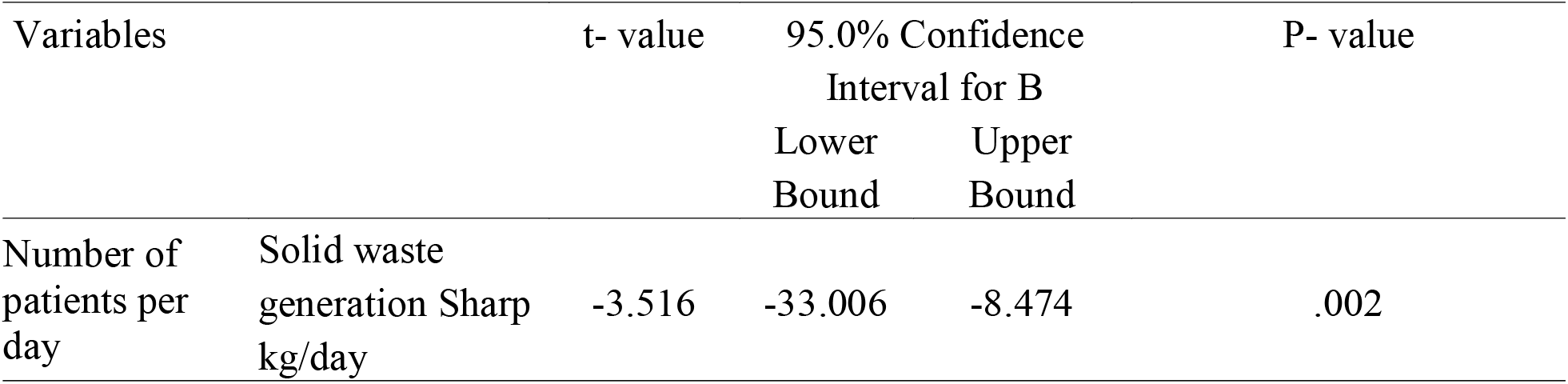
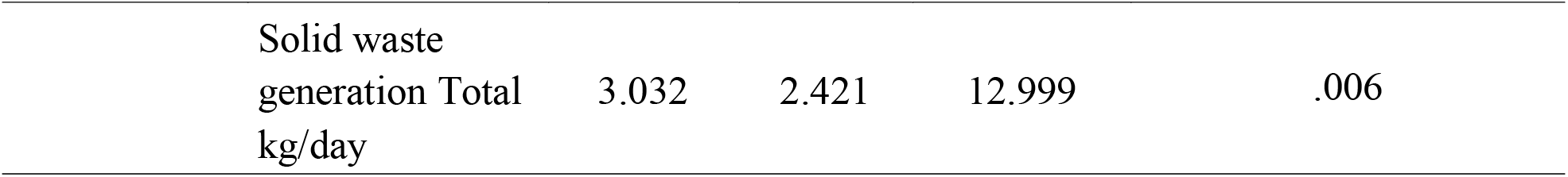
Linear regression analysis of the association of Number of patients per day with Total generation of solid biomedical waste in medical laboratories

Waste related to coronavirus pandemic is found to have a significant positive correlation with waste segregation practice (r=0.51, p=0.009) and availability of color coding (r=0.431, p=0.032). Table 7.

**Table 7:** Bivariate correlation analysis of waste of coronavirus pandemic with waste segregation practice and availability of color coding

| Variables |  | Waste segregation practice | Color coding |
| --- | --- | --- | --- |
| Waste of coronavirus pandemic | Pearson Correlation coefficient | .510 | .431 |
|  | P-value | .009 | .032 |

## Discussion

Estimating the biomedical waste generation rate is essential for health facilities to design and implement a better management system. This study has found the mean daily generation rate of biomedical waste in medical laboratories in health care facilities of Addis Ababa was 4.9 ± 3.13 kg/day per medical laboratory, of which 3.86 ± 2.66 kg/day was hazardous waste and 1.10 ± 0.853 kg/day was a sharp.. A study conducted in Adama city, Ethiopia revealed a mean daily generation of 4.46 ± 0.45kg/day per health facility[19].

The incremental waste generation rate in the current study might be because the study was conducted during COVID-19 pandemic which increases the waste generation rate due to extra precaution measures taken in health facilities. More over, medical laboratories generate a tremendous amount of weight-bearing biomedical wastes like glasses, tubes, sharps, and others[6]. The biomedical waste generation rate depends on the patient flow in the health care facility. A study in health care facilities in Addis Ababa found that there is a positive linear relationship between the number of patients and the generation rate of biomedical wastes[20]. This study also showed that there is a significant association between the number of patients getting medical laboratory service per day with the biomedical waste generation rate, P= 0.006. This indicates as the workload of the laboratory increases the waste generated in that laboratory as well increases.

Biomedical wastes generated in health care facilities including medical laboratories are dangerous health and environmental concerns which need proper management based on international and national guidelines. This study has found that currently, there are three guidelines regarding health care waste management in Ethiopia and 63% of laboratory professionals and 81.3% of waste handlers knew about the guidelines though 54.1% of them did indicate that the guidelines are not strong enough for implementing a proper practice. This result is consistent with a systematic review in Ethiopia which revealed that the regulations in Ethiopia are not updated and lack compliance with their implementation[16]. Another study in Ethiopia showed that the availability of health care waste management guidelines provides a particular quality to proper waste handling and management[21]. This study also found that knowledge about biomedical waste management policy and guidelines are significantly associated with the proper practice of management.

Training is found to be one of the essential components of effective biomedical waste management. This study found that 50.3% of study participants received training about biomedical waste management and as it had significant association with knowledge and proper practice of biomedical waste management is observed. This result was better than a finding from Debre Markos, the northwestern part of Ethiopia in which only 30.9% of the study participants were trained in waste management but did not comply with the national and international requirements[22]. Also, the far better result of only 2.9% of laboratory professionals were trained in a study in India[17].

The stuy revealed a great improvement that 96% of study participants confirmed medical laboratories in health care facilities used color coding based segregation in a container with a biohazard symbol. On the contrary, the systematic review mentioned the very limited waste segregation practice[16]. This could be since there is increase in the provision of training and increased awareness of laboratory managers. Additionally, the coronavirus pandemic had made professionals conscious to segregate waste related to the pandemic as 88% of them separately dispose of in infectious waste collection bin with cover.

In this study, 18.2% of participants encountered physical injuries like needle stick injury and sharp injury, and 15.6% of them occurred while handling biomedical waste which is consistent with the study in health facilities of Gondor town[9]. About 90.6% of the study participants of this study revealed as they use personal protective equipments for biomedical waste handling. This result is more or less similar to the study in Debre Markos which revealed 97% of study participants always use PPE while they are handling biomedical wastes[11].

Most medical laboratories in health facilities transport biomedical wastes using closed containers, though 39 % of them used open containers as another study in Addis Ababa found that open containers are used for transporting from collection site to storage and treatment[20]. The present study revealed that 56% had a waste storage area and 14.3% are secured. Conversely, out of the facilities that had a storage area 85.7% are open and not protected and 44% of the facilities had no storage area. A study in Ethiopia revealed a slightly different result which indicates 40% of health facilities stored their biomedical waste in an unprotected environment. The result of the study indicates health care waste storage practice is becoming poorer imposing health care professionals and the environment a great danger[11].

## Limitation of the study

Despite uncovering the generation rate and management practice of biomedical wastes from clinical laboratories in Ethiopia which produce dangerous infectious wastes and data is scarce, the study was limited only to measuring solid biomedical wastes. Due to financial and other inadequacies like lack of materials for measuring, the study could not include the generation rate and management of liquid biomedical waste management. Hence, we recommend further studies regarding the generation rate and management sytem of liquid biomedical waste in helath facilities of Addis Ababa Ethiopia for better understanding of the burden and identifiyig of gaps which would be inputs to design intervention mechanisms.

## Conclusion

The study revealed that medical laboratories generate a higher amount of biomedical waste per day which is significantly associated with the number of patients getting laboratory service. There is a gap of knowledge of policies and guidelines of biomedical waste management, common types, components, and infectious microorganisms related to biomedical wastes and their management. The provision of training was found to have a significant association with the proper knowledge and practice of biomedical waste management. This study indicated that there was a proper segregation practice, better waste storage and treatment practice for hazardous and non-hazardous waste. Conversely, inadequate, and inappropriate transportation tools were observed and open burning pits were commonly used for final waste disposal. Frequent training, making adjustments, and strengthening the implementation of the current national policy and guidelines of biomedical waste management in Ethiopia and more attention from laboratory and facility managers is very mandatory.

## Data Availability

All data are fully available without restriction and
all relevant data are within the manuscript and its Supporting Information files

## Acknowledgment

We would like to acknowledge Addis Ababa University for giving us the opportunity and financing the research. Our gratitude also goes to all data collectors and health facility mangers for their unreserved cooperation. Last but not least, we are indebted to the study participants without whom this study would not be realized.

